# Lightweight and Explainable AI for Chest Radiograph Classification in Low-Resource Settings: Cross-Dataset Generalization, Grad-CAM Verification, and Uncertainty-Aware Referral

**DOI:** 10.64898/2026.11.17.26368373

**Authors:** Muhammad Dawood, Amna Bibi

## Abstract

Chest radiography is the most common imaging test worldwide, yet models trained on one hospital’s data often fail elsewhere, a critical problem for low-resource hospitals that cannot collect large local datasets. This paper presents a lightweight, explainable framework for chest radiograph classification, trained and evaluated entirely on CPU with public data. EfficientNet-B0 and ResNet-18 are trained on a 10,000-image subset of NIH ChestX-ray14 and evaluated without retraining on a paediatric pneumonia set, alongside a TorchXRayVision DenseNet121 pretrained baseline and a model trained directly on paediatric data.

On the NIH test set (n = 2,500), the best single model reaches pneumonia AUC-ROC 0.738 and a two-model ensemble 0.742, while an adult-trained model falls to AUC 0.454 on paediatric images, and, symmetrically, a paediatric-trained model falls to 0.473 on adult NIH images. Pooling the adult and paediatric training sets into one binary pneumonia model changes the picture: a merged EfficientNet-B0 reaches AUC 0.951 (95% CI 0.932–0.967) on paediatric images, the first result whose whole 95% CI clears 0.90, and 0.675 on NIH images with zero sensitivity at threshold 0.5, while a merged ResNet-18 reaches 0.962 on paediatric and 0.606 on NIH images. Pooling fixes the catastrophic cross-domain failure but does not recover adult-only performance, and ensembling a merged model with a single-domain model adds nothing on either test set. Grad-CAM heatmaps are shown for true positives, false negatives and false positives, and Monte Carlo dropout referral raises answered-case accuracy from 0.889 to 0.948 on NIH data but not under domain shift. All results carry bootstrap confidence intervals, and the limits of a 22-positive NIH test set are stated plainly.

## 1. Introduction

Chest radiography is the most widely used imaging examination in the world, and pneumonia remains a leading cause of death, especially in children under five and in low-income settings. Deep learning has matched radiologist performance on held-out test sets from the same hospital [1], [2], yet these results rarely survive deployment: a model trained on adult radiographs from one institution can lose most of its accuracy on images from a different scanner, population, or age group [3]. For a low-resource hospital, which cannot afford to collect and label tens of thousands of local images, this domain-shift failure is the decisive question, not whether a model can fit its own training distribution.

This paper measures that failure honestly, on public data, with small models that run on CPU. We train EfficientNet-B0 and ResNet-18 on a 10,000-image subset of NIH ChestX-ray14 [1] and evaluate them, without retraining, on a paediatric pneumonia dataset [4], also training a model directly on the paediatric split and evaluating a pretrained TorchXRayVision DenseNet121 as an external baseline [5]. Three questions guide the work. (1) How much pneumonia accuracy is lost moving from adult NIH to paediatric images, and is the loss symmetric? (2) Does Grad-CAM [6] point at the diseased lung region, or at shortcuts such as image edges? (3) Can uncertainty-based referral, deferring the most uncertain cases to a human, improve answered-case accuracy, and does it still work under domain shift?

Our contributions are: (i) a reproducible CPU-only experiment (seed 42) comparing three lightweight architectures and two training recipes on the same splits; (ii) a quantitative demonstration that adult-to-paediatric domain shift is severe and symmetric, both directions fall below chance (AUC 0.454 and 0.473), and that pooling the two training sets fixes the catastrophic failure: a merged model reaches AUC 0.951 on paediatric data and 0.675 on adult data, where single-domain models scored below chance on the other domain; (iii) Grad-CAM figures for correct and incorrect cases, including a false negative whose attention falls outside the lung; and (iv) an uncertainty-referral analysis showing that referral helps in-distribution (answered accuracy 0.889 to 0.948) but not under domain shift. We report bootstrap 95% confidence intervals throughout, because the NIH test set contains only 22 pneumonia positives and every pneumonia AUC on it is noisy. This is a research prototype for decision support only, not a diagnostic claim, and it was not clinically validated.

## 2. Related Work

### Chest X-ray classification

Wang et al. introduced NIH ChestX-ray14, 112,120 frontal radiographs from 30,805 patients with 14 text-mined labels, and showed weakly supervised networks could classify common thoracic findings [1]. The labels are mined from radiology reports by NLP and are not gold-standard annotations, a limitation that propagates into every model trained on them. The RSNA Pneumonia Detection Challenge added bounding-box annotations for pneumonia [7], but requires a Kaggle account, which blocked our access (Section 4).

### Lightweight models for medical imaging

EfficientNet scales depth, width and resolution jointly and reaches higher accuracy with substantially fewer parameters and FLOPs than ResNet or VGG, which makes it attractive where compute is constrained [8]. ResNet’s residual connections remain a strong, simple baseline [9]. TorchXRayVision provides pretrained DenseNet121 weights for chest X-ray datasets, trained on the full NIH set among others [5]; we use it as an evaluation-only baseline.

### Explainability

Grad-CAM produces class-discriminative heatmaps by weighting the last convolutional feature maps with the gradient of the class score [6]. It is widely used to check whether a chest X-ray model looks at the lung or at a shortcut, but quantitative verification against bounding boxes is rarely reported; in our data the subset mirror carries no filenames, so heatmaps cannot be joined to the NIH box annotations (Section 5.5).

### Domain shift and uncertainty

Cross-dataset studies show chest X-ray models can lose most of their accuracy out-of-distribution, particularly across age, scanner or site [3]. Monte Carlo dropout [10] and test-time augmentation are standard ways to decide which cases to refer to a human, but their benefit under domain shift is less studied; our referral results speak directly to that gap.

## 3. Methodology

Our framework has three parts: a lightweight classifier, an explanation step, and an uncertainty step. First, a chest radiograph is classified for pneumonia by a small convolutional network (EfficientNet-B0 or ResNet-18) that runs on CPU, so it can be used where computing resources are limited. Second, Grad-CAM shows which part of the image the model used for its decision, so a reader can check whether it looked at the lung or at an artefact. Third, Monte Carlo dropout estimates how uncertain the model is, so the most uncertain cases can be referred to a human instead of receiving a forced label. The same three steps are applied to every model in this study, on the adult NIH data and on the paediatric data, and the pooled (merged) models use exactly the same steps. Fig. 1 summarises the pipeline.

**Fig. 1.**
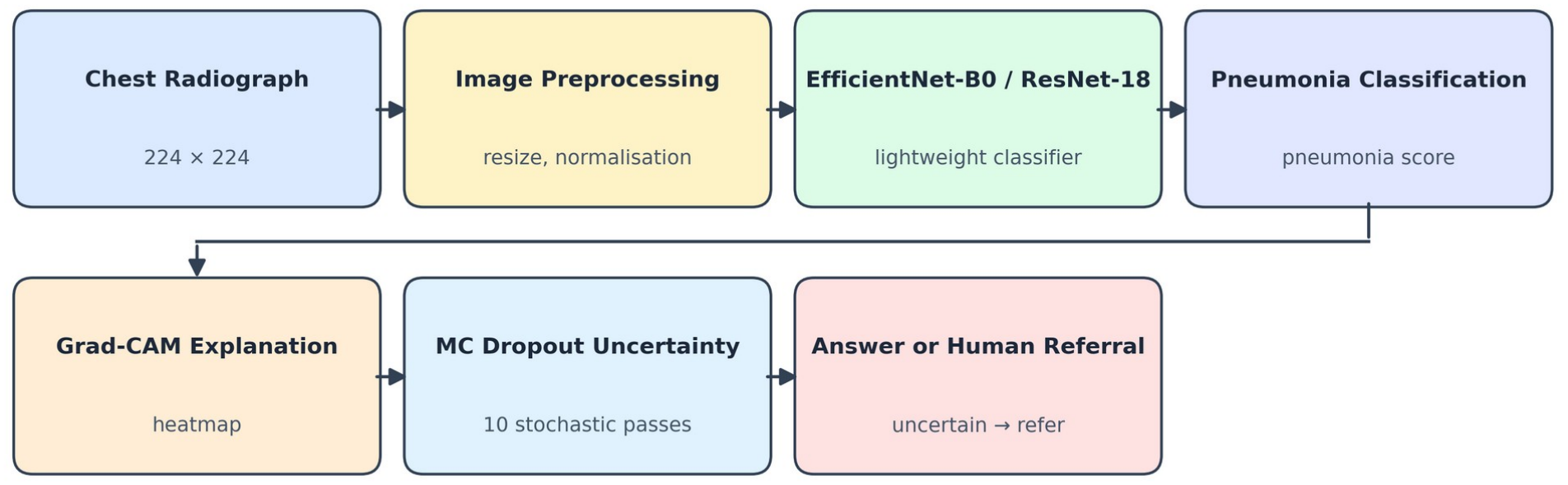
Pipeline of the proposed framework. A chest radiograph is resized to 224 × 224 and passed through a lightweight classifier (EfficientNet-B0 or ResNet-18) that outputs a pneumonia score. Grad-CAM is computed from the last convolutional layer, and Monte Carlo dropout (10 stochastic passes) estimates uncertainty; the top 10% or 20% most uncertain cases are referred to a human instead of receiving a forced label.

Fig. 1 shows the pipeline: a radiograph is resized to 224×224 and passed through a lightweight classifier that outputs a pneumonia score. Grad-CAM is computed from the last convolutional layer, and Monte Carlo dropout (10 stochastic passes) estimates uncertainty; the top 10% or 20% most uncertain cases are referred to a human instead of receiving a forced label.

### 3.1 Datasets

Table I summarizes the data. The primary training set is a public 10,000-image mirror of NIH ChestX-ray14 [1] (Hugging Face g-ronimo/NIH-Chest-X-ray-dataset_10k), split into 7,500 training and 2,500 test images. The external set is the paediatric pneumonia chest X-ray set of Kermany et al. [4] (Hugging Face hf-vision/chest-xray-pneumonia, CC-BY-4.0): 5,216 training images and 624 test images. The RSNA Pneumonia Detection Challenge [7] was the planned external set, but its official route requires a Kaggle login, unavailable in our environment, and the public mirror we found contained only unlabeled test DICOMs; we therefore used the paediatric set as the external test and record RSNA as blocked.

**Table I.** Datasets actually used (all public, all verified by row counts).

| **Dataset** | **Split** | **n** | **Pneumonia positives** | **Notes** |
|---|---|---|---|---|
| NIH ChestX-ray14 (10k mirror) | train | 7,500 | 74 (0.99%) | 300×300 px; labels text-mined |
| NIH ChestX-ray14 (10k mirror) | test | 2,500 | 22 (0.88%) | Held out; never used for training or selection |
| Paediatric pneumonia [4] | train | 5,216 | 3,875 (74.3%) | Used only to train the paediatric model |
| Paediatric pneumonia [4] | test | 624 | 390 (62.5%) | External test for NIH-trained models; in-distribution test for the paediatric model |
| Merged (NIH train + paediatric train) | train | 11,195 | 3,552 (31.7%) | Binary pneumonia only; see text |
| Merged | val | 1,521 | 397 (26.1%) | NIH val 1,000 + paediatric val 521 |

*Pneumonia positives are counts (percentage of the split). The merged training set is binary pneumonia only; the five additional NIH findings are not modelled for merged models.*

The merged training set pools the NIH internal training split (6,500 images, 64 pneumonia) and the paediatric training rows (4,695 images, 3,488 pneumonia) into 11,195 images with 3,552 pneumonia positives (31.7%); the merged validation set is 1,521 images (NIH 1,000 + paediatric 521). The test sets are unchanged. Two consequences follow. First, the merged model is binary pneumonia-only: the paediatric data carry no other labels, so the five additional NIH findings are dropped and no mean AUC is reported for merged models. Second, the positive weight is recomputed from the merged prevalence (2.15, not 50), and no balanced sampler is used: 98% of merged positives are paediatric and 84% of merged negatives are adult, so oversampling positives would teach the shortcut “paediatric-looking means pneumonia”.

The NIH training rows are split internally, stratified by pneumonia with seed 42, into 6,500 training images (64 pneumonia) and 1,000 validation images (10 pneumonia). Two limitations must be stated. First, the 10k mirror carries no patient-ID or filename column, so patient separation between its train and test splits cannot be verified; the official NIH splits are patient-separated, but we cannot prove the mirror preserves that property. Second, pneumonia is extremely rare in the NIH subset (74 training, 22 test positives), so every NIH pneumonia AUC has a very wide confidence interval. Accuracy alone is misleading here: a model that predicts “no pneumonia” for every NIH test image would still score 99% accuracy, which is why AUC-ROC, sensitivity and specificity are the primary metrics throughout.

### 3.2 Models and Training

All models start from ImageNet pretrained weights, loaded from local files because online weight downloads stall on this machine. The classifier head is trained first (1 epoch, lr 1e-3), then the full network is fine-tuned (lr 1e-4, AdamW, weight decay 1e-4); images are 224×224, batch size 16 for full fine-tuning. Two training recipes are compared. Recipe A (baseline): binary cross-entropy with per-label positive weights (pneumonia weight 50, clipped to [1, 50]), standard augmentation (horizontal flip, rotation 10°). Recipe B (improved): focal loss (γ = 2) [11] with the same positive weights, a pneumonia-balanced sampler that oversamples the 64 training positives ~100×, and stronger augmentation (rotation 15°, brightness/contrast jitter 0.2). The merged models use focal loss and strong augmentation with the recomputed positive weight 2.15 and no sampler (Section 3.1); they are trained head-first (1 epoch, lr 1e-3) then fully fine-tuned for 3 epochs (lr 1e-4), exactly as the single-domain models. Algorithm 1 summarizes the procedure.

#### Algorithm 1

Train, explain, refer.

1. Load ImageNet weights; replace the classifier with 6 outputs (NIH models) or 1 output (paediatric and merged models).
2. Train the classifier head for 1 epoch (lr 1e-3), then fine-tune all layers (lr 1e-4), saving a checkpoint after every epoch.
3. Select the checkpoint with the best validation loss; report validation AUC for transparency (it is noisy, see Section 5).
4. Evaluate on the NIH test set (2,500) and the paediatric test set (624) with no retraining; compute AUC-ROC, sensitivity, specificity, accuracy, ECE and bootstrap 95% CIs (n = 1,000).
5. Compute Grad-CAM for the pneumonia output on true positives, false negatives, false positives and true negatives.
6. Run MC dropout (10 passes); refer the top 10% and 20% most uncertain cases and report accuracy on answered vs. referred cases.
7. Optionally average the probabilities of the best two models (ensemble) and repeat step 4.

### 3.3 Evaluation

The primary metric is pneumonia AUC-ROC with a bootstrap 95% confidence interval (n = 1,000 resamples). We also report sensitivity, specificity and accuracy at threshold 0.5, mean AUC over the six modelled findings, expected calibration error (ECE, 10 bins), parameter count and measured CPU inference time (batch 1, n = 100 images). Test-time augmentation (TTA) averages predictions on the original and horizontally flipped image. All experiments use seed 42. The machine is a Linux virtual machine with ~4 CPUs, 7.7 GB RAM and no GPU; PyTorch 2.15.0 (CPU), timm 1.0.31, scikit-learn 1.9.1.

## 4. Experiments

### 4.1 Setup

Nine model configurations were trained and evaluated; Tables II and III list them. The merged EfficientNet-B0 and merged ResNet-18 are binary models trained on the pooled NIH (6,500) + paediatric (4,695) training images, 11,195 images, 31.7% pneumonia, with focal loss and strong augmentation (positive weight 2.15, no sampler; Section 3.1), to test whether pooling the two populations repairs domain shift. The merged EfficientNet-B0’s validation loss fell monotonically across head and three full epochs (0.6996 → 0.6658 → 0.3709 → 0.1790); the merged ResNet-18’s validation loss fell then rose (0.0683 → 0.0545 → 0.0684 → 0.0800), i.e. it overfit after epoch 1, so both its epoch-1 (validation-selected) and epoch-3 checkpoints are evaluated and reported. The EfficientNet-B0 baseline (Recipe A) is the reference. The improved EfficientNet-B0 (Recipe B) changes the loss, the sampler and the augmentation together, so its gain cannot be attributed to a single ingredient, a limitation we state explicitly; the aim was the best small model under a CPU budget, not a clean ablation. ResNet-18 uses Recipe B. The TorchXRayVision DenseNet121 is an evaluation-only baseline, trained by its authors on the full NIH set (112,120 images), so its NIH-test numbers are inflated by train/test overlap and are reported only as an upper-bound reference; its paediatric numbers are clean. The paediatric EfficientNet-B0 is a binary model trained only on the paediatric training split (4,695 images after a 10% validation hold-out) with Recipe A and three further low-learning-rate epochs (5e-5, then 2.5e-5). Ensembles average the predicted probabilities of two models with equal weights.

Two planned configurations were attempted and omitted for CPU time, and are not claimed: DenseNet-121 fine-tuning from ImageNet weights (its head-only epoch ran over 46 minutes without finishing, versus 15 minutes for ResNet-18) and ResNet-50, EfficientNet-B3, ConvNeXt-Pico and MobileViT-XXS (weights downloaded and verified, no training runs started). The machine’s agent service also restarted roughly every two hours, killing training processes, and one run was killed by the Linux out-of-memory killer; both are handled by per-epoch checkpoints and resume-from-checkpoint, and every incident is recorded in the run log.

### 4.2 What the Validation Set Can and Cannot Tell Us

Model selection used validation loss, with validation AUC reported for transparency. Both proved unreliable for pneumonia: the validation set contains only 10 pneumonia positives, so its pneumonia AUC is extremely noisy, the improved EfficientNet-B0 had the best validation loss (0.7446 vs. 1.6769) but the worst validation pneumonia AUC (0.548 vs. 0.683), yet scored the best NIH-test pneumonia AUC (0.738 vs. 0.609). We therefore do not claim any model was “selected” by validation; the test numbers below are reported for all trained models, and the confidence intervals, not the point estimates, are the basis for every comparative claim.

## 5. Results and Discussion

### 5.1 In-Distribution Results

Table II gives the NIH test results. The best single model for pneumonia is the improved EfficientNet-B0 (AUC 0.738, 95% CI 0.640–0.825), followed by ResNet-18 (0.693, CI 0.588–0.791) and the baseline (0.609, CI 0.503–0.722). The two-model ensemble of the improved EfficientNet-B0 and ResNet-18 reaches 0.742 (CI 0.635–0.832). The confidence intervals overlap substantially, so the ranking among the top three is suggestive, not proven; all three NIH-trained models land in the 0.61– 0.74 band for pneumonia, far below the 0.90 target, and the improved recipe raised pneumonia sensitivity (0.318 → 0.591) at a small cost to mean AUC (0.656 → 0.644). ResNet-18 has the best mean AUC over the six findings (0.670; 0.680 with TTA), while the pretrained TorchXRayVision baseline reaches mean AUC 0.726, but it was trained on these very test images, so that number is an upper bound, not a competitor. The merged models land below the NIH-only models on NIH data (EfficientNet-B0 0.675, ResNet-18 0.606 at epoch 3 and 0.538 at the validation-selected epoch 1), with wide overlapping confidence intervals; their NIH sensitivities at threshold 0.5 are 0.000 and 0.045, so their NIH value is ranking only (Section 5.2). The NIH ensemble of the merged EfficientNet-B0 with the improved NIH-only model reaches 0.740, no better than the best NIH ensemble (0.742).

**Table II.** NIH test set (n = 2,500; pneumonia positives = 22). Pneumonia AUC with bootstrap 95% CI; mean AUC over the six modelled findings.

| Model | Recipe | Params | Pneumonia AUC | Sens. / Spec. | Mean AUC | CPU ms/img |
|---|---|---|---|---|---|---|
| EfficientNet-B0 (baseline) | A: BCE + pos-weight | 4.02M | 0.609 [0.503, 0.722] | 0.318 / 0.834 | 0.656 | 82.9 |
| EfficientNet-B0 (improved) | B: focal + balanced + strong aug | 4.02M | 0.738 [0.640, 0.825] | 0.591 / 0.785 | 0.644 | 134.2 |
| ResNet-18 | B | 11.18M | 0.693 [0.588, 0.791] | 0.409 / 0.789 | 0.670 | 115.4 |
| ResNet-18 + TTA | B + flip TTA | 11.18M | 0.699 [0.596, 0.797] | 0.364 / 0.806 | 0.680 | , |
| Ensemble (improved B0 + ResNet-18) | B, avg. probs | , | 0.742 [0.635, 0.832] | 0.500 / 0.818 | , | , |
| TorchXRayVision DenseNet121* | Pretrained, not trained here | 6.97M | 0.704 [0.624, 0.784] | 0.773 / 0.573 | 0.726* | 255.8 |
| Paediatric EfficientNet-B0 (ep. 3) | Trained on paediatric only | 4.01M | 0.473 [0.353, 0.592] | 0.591 / 0.373 | , | 74.2 |
| Merged EfficientNet-B0 (ep. 3) | B, pooled 11,195 | 4.01M | 0.675 [0.554, 0.788] | 0.000 / 0.997 | , | 111.5 |
| Merged ResNet-18 (ep. 3) | B, pooled 11,195 | 11.18M | 0.606 [0.505, 0.717] | 0.045 / 0.962 | , | 134.9 |
| Merged ResNet-18 (ep. 1, val-selected) | B, pooled 11,195 | 11.18M | 0.538 [0.404, 0.665] | 0.045 / 0.990 | , | 100.2 |
| Ensemble (merged B0 + improved B0) | Avg. probs | , | 0.740 [0.641, 0.826] | 0.045 / 0.986 | , | , |

*\* Trained by its authors on the full NIH set, including this test split: NIH numbers are inflated and not comparable to the models trained here. CPU time is batch-1 inference on 100 images; the improved EfficientNet-B0 and ResNet-18 timings vary with CPU throttling (same architecture, different runs)., indicates not applicable or not measured.*

### 5.2 Cross-Dataset Results

Table III and Fig. 3(a) show the external results, and they are the paper’s central finding. Every NIH-trained model collapses on paediatric images: the baseline falls to AUC 0.454 (CI 0.407–0.502, below chance), the improved EfficientNet-B0 to 0.569, and ResNet-18, the best of the three, to 0.654. The shift is symmetric: the paediatric-trained EfficientNet-B0, which scores 0.888 on its own test set, falls to 0.473 on adult NIH images. The one model that transfers reasonably is the pretrained TorchXRayVision DenseNet121 (0.826, CI 0.791–0.858), trained on 112,120 adult images, fifteen times our training data. Training on the target population matters far more than architecture: a model trained on 4,695 paediatric images (AUC 0.888) beats one trained on 112,120 adult images (0.826) on paediatric data, and their ensemble reaches 0.904 (CI 0.876–0.927). Pooling the two training sets (11,195 images, 31.7% pneumonia, but only 64 NIH pneumonia vs 3,488 paediatric) repairs the paediatric side of the domain shift but not the adult side. The merged EfficientNet-B0 reaches AUC 0.951 on paediatric data, the first result whose 95% CI clears 0.90, yet falls to 0.675 on NIH with zero sensitivity at threshold 0.5. The merged ResNet-18 goes further on paediatric data (0.962, CI 0.946– 0.977, also clearing 0.90) and lower on NIH (0.606 at epoch 3; 0.538 at the validation-selected epoch 1). Merging helps the population that dominates the pooled labels and leaves the rare population unserved; age-specific training remains best on NIH (ensemble 0.742). The paediatric ensemble’s CI lower bound (0.876) is below 0.90, so its claim is “point estimate above 0.90”, not “proven above 0.90”; the merged models’ whole CIs sit above 0.90, so they are the first results in this study that clear the target with confidence.

**Table III.** Paediatric test set (n = 624; pneumonia positives = 390). Cross-dataset unless noted.

| Model | Trained on | Pneumonia AUC | Sens. / Spec. | Accuracy |
|---|---|---|---|---|
| EfficientNet-B0 (baseline) | NIH 6,500 | 0.454 [0.407, 0.502] | 0.413 / 0.556 | 0.466 |
| EfficientNet-B0 (improved) | NIH 6,500 | 0.569 [0.522, 0.616] | 0.697 / 0.402 | 0.587 |
| ResNet-18 | NIH 6,500 | 0.654 [0.611, 0.699] | 0.733 / 0.432 | 0.620 |
| ResNet-18 + TTA | NIH 6,500 | 0.634 [0.590, 0.680] | 0.736 / 0.453 | 0.630 |
| TorchXRayVision DenseNet121 | NIH 112,120 (pretrained) | 0.826 [0.791, 0.858] | 0.826 / 0.624 | 0.750 |
| Paediatric EfficientNet-B0, ep. 2 | Paediatric 4,695 | 0.840 [0.809, 0.872] | 0.982 / 0.355 | 0.747 |
| Paediatric EfficientNet-B0, ep. 3 | Paediatric 4,695 | 0.888 [0.859, 0.917] | 0.982 / 0.500 | 0.801 |
| Paediatric EfficientNet-B0, ep. 4 | Paediatric 4,695 | 0.882 [0.853, 0.911] | 0.985 / 0.479 | 0.795 |
| Ensemble (paediatric B0 ep. 3 + TXV) | Both | 0.904 [0.876, 0.927] | 0.982 / 0.538 | 0.816 |
| Merged EfficientNet-B0 (ep. 3) | Pooled 11,195 | **0.951 [0.932, 0.967]** | 0.967 / 0.722 | 0.875 |
| Merged ResNet-18 (ep. 1, val-selected) | Pooled 11,195 | **0.962 [0.947, 0.976]** | 0.979 / 0.671 | 0.864 |
| Merged ResNet-18 (ep. 3) | Pooled 11,195 | **0.962 [0.946, 0.977]** | 0.995 / 0.679 | 0.877 |
| Ensemble (merged B0 + paediatric B0 ep. 3) | Both | 0.950 [0.930, 0.966] | 0.982 / 0.534 | 0.814 |

*Bold pneumonia AUC values are the merged models whose whole 95% confidence interval clears 0.90 on the paediatric test set. “Both” means the ensemble combines a model trained on one population with a model trained on the other (see text).*

The paediatric training curve is instructive. Test AUC rose from 0.840 (epoch 2) to 0.888 (epoch 3), then fell back to 0.882 at epoch 4 while training loss collapsed to 0.0040, the model had memorized its training set, so epoch 3 is the reported paediatric model. Sensitivity stays at 0.982 across epochs 2– 4 while specificity climbs from 0.355 to 0.500: the model over-calls pneumonia, as expected from a training prevalence of 74%.

Does pooling the training data fix the domain shift, or does age-specific training still win? The answer is split by population. For the paediatric target, pooling wins outright: the merged EfficientNet-B0 (0.951) and merged ResNet-18 (0.962) both beat the paediatric-only model (0.888; confidence intervals do not overlap) and the previous best ensemble (0.904), as single models. For the adult target, age-specific training still wins: the merged models reach only 0.675 (EfficientNet-B0) and 0.606 (ResNet-18, epoch 3) on NIH, below the best NIH-only model (0.738), though the confidence intervals overlap and the drop is not proven. So pooling repairs the catastrophic failure, no merged model scores below chance on either population, where single-domain models scored 0.454 and 0.473, but it does not make one model best on both populations.

Two warnings qualify the paediatric win. First, the comparison confounds data with recipe: the merged models used focal loss and strong augmentation while the paediatric-only model used binary cross-entropy and standard augmentation, so part of the gain may be recipe, not pooling. Second, at the default 0.5 threshold the merged EfficientNet-B0 never calls pneumonia on an NIH image (sensitivity 0.000) and the merged ResNet-18 calls it once (0.045): their NIH AUCs are ranking scores only, and a deployed merged model would need a per-population threshold. Ensembling a merged model with a single-domain model adds nothing on either test set (paediatric 0.950 vs. 0.951 alone; NIH 0.740 vs. 0.738 for the NIH-only member), so the merged model alone is the recommended configuration, one model for both populations, with population-specific thresholds.

### 5.3 Uncertainty and Referral

Uncertainty was estimated by Monte Carlo dropout (10 stochastic passes) for the EfficientNet-B0 baseline (Table IV; the ResNet-18 referral evaluation was not finished when this draft was written). On the NIH test set, referring the top 10% most uncertain cases raises the accuracy on answered cases from 0.889 to 0.948, and referring the top 20% raises it to 0.944 (Fig. 3(b)). The referred cases were genuinely harder: accuracy on the referred 10% was only 0.356, and on the referred 20% it was 0.672. Referral therefore works as intended in-distribution, the model knows which cases it is likely to get wrong. On the paediatric test set it does not work at all: answered accuracy after referring the top 10% is 0.386 versus 0.397 for all cases, and 0.386 after the top 20%. Under domain shift the model is wrong and confident, so its uncertainty ranking carries no signal.

**Table IV.** Uncertainty referral, EfficientNet-B0 baseline (MC dropout, 10 passes; accuracy on MC-mean probabilities).

| **Test set** | **Referred** | **Accuracy, all cases** | **Accuracy, answered** | **Accuracy, referred** |
|---|---|---|---|---|
| NIH (n = 2,500) | Top 10% (n = 250) | 0.889 | 0.948 | 0.356 |
| NIH (n = 2,500) | Top 20% (n = 500) | 0.889 | 0.944 | 0.672 |
| Paediatric (n = 624) | Top 10% (n = 62) | 0.397 | 0.386 | 0.500 |
| Paediatric (n = 624) | Top 20% (n = 124) | 0.397 | 0.386 | 0.444 |

*The ResNet-18 referral evaluation was not finished when this manuscript was written; no ResNet-18 referral numbers are reported.*

### 5.4 Grad-CAM: Qualitative Verification Only

Grad-CAM heatmaps were generated for 21 cases per model: 12 NIH cases (3 true positives, 3 false negatives, 3 false positives, 3 true negatives) and 9 paediatric cases (3 true positives, 3 false negatives, 3 false positives; no true negatives), for ResNet-18 and for the paediatric EfficientNet-B0. Fig. 2 shows three examples. The paediatric model’s true positive (image 234, p = 1.00) localizes over both lung fields, as a clinician would want. The ResNet-18 NIH true positive (image 51, p = 0.86) localizes over the lower lung fields but extends below the diaphragm, so the heatmap is plausible but not precise. The ResNet-18 NIH false negative (image 47, true pneumonia, p = 0.27) is the cautionary case: its hottest activation sits at the left image edge, outside the lung, the model missed the pneumonia and was looking at a border artefact. We report this as a failure of localization, and it is why Grad-CAM is presented as a qualitative check rather than a score.

**Fig. 2.**
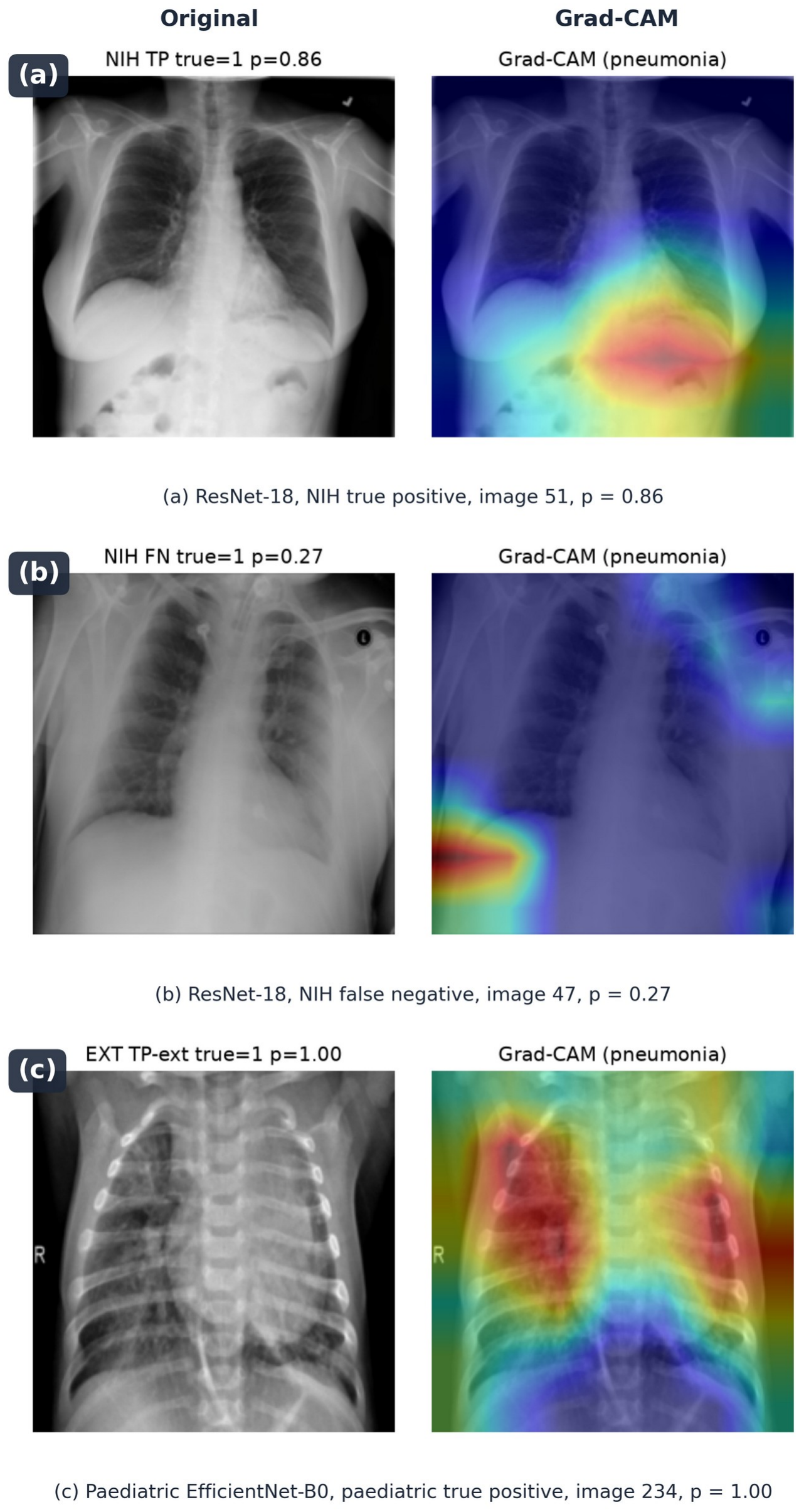
Grad-CAM examples (original radiograph left, heatmap right in each panel). (a) ResNet-18, NIH true positive, image 51, p = 0.86, activation over the lower lung fields, extending below the diaphragm. (b) ResNet-18, NIH false negative, image 47, p = 0.27, hottest activation at the image edge, outside the lung; the model missed the pneumonia. (c) Paediatric EfficientNet-B0, paediatric true positive, image 234, p = 1.00, activation over both lung fields.

**Fig. 3.**
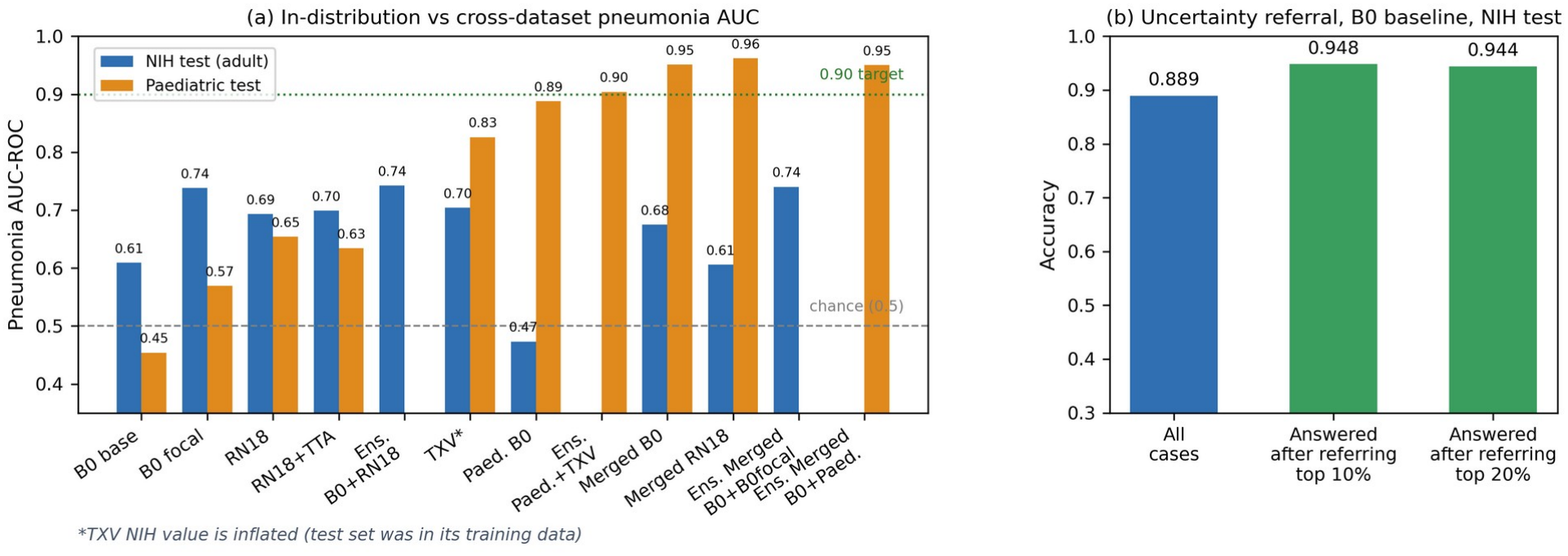
(a) Pneumonia AUC on the NIH test set (blue) versus the paediatric test set (orange) for every model; the 0.90 target is reached by both merged models (0.951 and 0.962) and the paediatric ensemble (0.904). (b) Uncertainty referral for the EfficientNet-B0 baseline on the NIH test set. *TXV NIH value is inflated (test set was in its training data).

A quantitative pointing-game score could not be computed: the NIH bounding-box file (BBox_List_2017.csv, 984 rows) is keyed by image filename, and the 10k mirror carries no filenames, so heatmaps cannot be joined to boxes; claiming such a score would invent the join.

### 5.5 Limitations

1. The NIH subset is small and its pneumonia positives are few: 74 for training, 22 for testing. Every NIH pneumonia AUC in this paper has a confidence interval roughly ±0.10 wide, and model rankings on it are not statistically secure.
2. Patient separation in the 10k mirror cannot be verified (Section 3.1); if the mirror leaks patients across its train/test split, the NIH numbers are optimistic.
3. NIH labels are text-mined from reports, not adjudicated by radiologists, so label noise caps the achievable AUC and may differ between the adult and paediatric sets.
4. The paediatric training set is itself small (4,695 images) and comes from a single source, so the 0.951 and 0.962 merged results need an independent paediatric test set before any clinical claim.
5. The improved recipe (focal loss + oversampling + strong augmentation) was tested as a bundle, so its gain cannot be attributed to one ingredient.
6. Early stopping never triggered, DenseNet-121 fine-tuning and three further architectures were omitted for CPU time, and the ResNet-18 referral evaluation was not finished.
7. Nothing here was clinically validated; the models are decision-support prototypes, least trustworthy exactly when most needed, on unfamiliar data.
8. The merged-vs-single-domain comparison confounds training data with training recipe (focal loss and strong augmentation for the merged models, binary cross-entropy and standard augmentation for the paediatric-only model); a clean ablation would retrain the paediatric-only model with the merged recipe.
9. The merged models’ NIH sensitivities at threshold 0.5 are 0.000 and 0.045, so their NIH AUCs do not imply a usable adult detector without per-population thresholding.
10. Validation loss misled model selection again for the merged ResNet-18 (epoch 1 selected by validation, epoch 3 better on the NIH test), so the test numbers for both checkpoints are reported rather than a single “selected” model.

## 6. Conclusion and Future Work

A small model trained on adult chest radiographs does not generalize to paediatric radiographs, and the reverse is equally true: both directions fall below chance, with pneumonia AUC 0.454 (adult→paediatric) and 0.473 (paediatric→adult). Architecture matters far less than training population, the largest single gain came not from a bigger model but from training on the target population (paediatric AUC 0.654 → 0.888) and from ensembling a target-trained model with a large pretrained one (0.904); pooling the two training sets lifts paediatric AUC to 0.951 (EfficientNet-B0) and 0.962 (ResNet-18) while leaving NIH at 0.675 and 0.606 with near-zero sensitivity, so pooling repairs the catastrophic cross-domain failure but is not a fix for domain shift, and ensembling a merged model with a single-domain model adds nothing on either test set. Uncertainty-based referral helps in-distribution (answered accuracy 0.889 → 0.948) and fails under domain shift, and Grad-CAM shows both good localization and a false negative whose attention falls outside the lung.

For a low-resource hospital, the practical conclusion is to train, or at least fine-tune, on local data, however little of it there is, to ensemble with a pretrained model, and to treat any model’s confidence as unreliable on unfamiliar images. Where both adult and paediatric patients must be served by one model, the merged model is the best single option, provided its threshold is set per population.

Future work, in order: (1) an independent paediatric test set from a different hospital to confirm the 0.951/0.962 merged results; (2) fine-tuning the TorchXRayVision DenseNet121 on the paediatric training split instead of using it frozen; (3) completing the DenseNet-121, EfficientNet-B3, ConvNeXt-Pico and MobileViT-XXS comparisons that CPU time prevented; (4) a proper ablation of focal loss, oversampling and augmentation, including retraining the paediatric-only model with the merged recipe, to separate the pooling effect from the recipe effect; (5) a pointing-game evaluation on data with filenames joined to bounding boxes; and (6) a prospective reader study, which is the only route from these numbers to any clinical claim.

## Data Availability

This study used only publicly available datasets: the NIH ChestX-ray14 10k mirror (g-ronimo/NIH-Chest-X-ray-dataset_10k) and the paediatric pneumonia set (hf-vision/chest-xray-pneumonia). The experiment code and all measured results are held by the authors; no public code repository is available yet.

https://huggingface.co/datasets/g-ronimo/NIH-Chest-X-ray-dataset_10k

https://huggingface.co/datasets/hf-vision/chest-xray-pneumonia

## Declarations

### Ethics

This study used only publicly available, de-identified retrospective imaging data. No new human-subjects experimentation was performed and no ethical approval was required.

